# SARS-CoV-2 infection control implementation based on sources of infection showing directions for three age groups in Japan

**DOI:** 10.1101/2021.01.14.21249637

**Authors:** Atsuko Hata, Junko Kurita, Eri Muso, Toshiro Katayama, Takahide Hata, Yasushi Ohkusa

## Abstract

**Background:** Some aspects of severe acute respiratory syndrome coronavirus 2 (SARS-CoV-2) transmission in children and adults remain unclear. This report describes different SARS-CoV-2 transmission patterns by age group in Japan.

**Methods and findings:** This retrospective observational case series study analyzed transmission patterns of real-time polymerase chain reaction (RT-PCR)-confirmed SARS-CoV-2 infections found by local health authorities and commercial laboratories during January 14 through July 31, 2020 in Japan. After ascertaining the infection source for every symptomatic case as clusters at households, daycare facilities, schools, hospitals and workplaces etc., their associated transmission patterns were analyzed. Identified cases were divided into three groups: underage, < 20; adults, 20–59; and elderly people 60 years old and older. The reproductive number (R)s of respective transmission directions found for the respective age groups were compared.

Of 26,986 total cases, 23,746 unknown cases were found, leaving 3,240 ascertained sources of infection (12.0%) comprising 125 (3.9%) underage, 2350 (72.5%) adult, and 765 (23.6%) elderly people. The respective Rs of underage infection sources directed to underage, adult, and elderly people were estimated respectively as 0.0415 (95% CI, 0.0138–0.0691), 1.11 (95% CI, 0.9171–1.3226), and 0.2811 (95% CI, 0.2074–0.3687). The respective Rs of adult infection source directed to underage, adult, and elderly people were estimated respectively as 0.0140 (95% CI, 0.0120–0.0162), 0.5392 (95% CI, 0.5236–0.5550), and 0.1135 (95% CI, 0.1074–0.1197). The respective Rs of elderly infection source directed to underage, adult, and elderly people were estimated as 0.065 (95% CI, 0.0039–0.0091), 0.3264 (95% CI, 0.3059–0.3474), and 0.3991 (95% CI, 0.3757–0.4229).

**Conclusions:** The main sources of SARS-CoV-2 infection were adults and elderly people. The R of underage people directed to adults was greater than 1 because of close familial contact but they were unlikely to become carriers transmitting SARS-CoV-2 because they accounted for a minority for transmissions. Apparently, SARS-CoV-2 was transmitted among adults and elderly people, suggesting that infection control of SARS-CoV-2 should be managed specifically by generation.

## Introduction

At the beginning of the spread of the novel coronavirus infection, isolation and life restrictions imposed on all age groups were necessary measures to prevent the spread of novel viruses of unknown identity, to ascertain the pathogenicity, and to avoid difficult conditions of medical care. However, the long-term negative socioeconomic effects of these measures have also been considerable [1]. Adverse effects on education and exercise restrictions, especially for minors, are unfathomable. Nationwide surveys investigating SARS-CoV-2 transmission patterns among children, adolescents, adults, and elderly people in Japan have probably brought about a new age-dependent isolation strategy without the spread of viral infection.

In Japan, since the first person infected with the novel coronavirus was identified on January 15, 2020, 4,314,979 confirmed cases of the novel coronavirus disease (COVID-19) had been recorded by December 27, 2020. The death toll had risen to 3,212 in Japan by December 27, 2020. Of 202,128 infection cases, 12,209 (6.0%) were of young people aged 10–19; 4,743 (2.3%) were of people younger than 10 years old on December 23, 2020 [2]. In response to the COVID-19 pandemic, numerous countries have implemented public health and social measures (PHSM), including school closure [3]. The Japanese government asked all schools and kindergartens to close from March 2, 2020 to prevent the spread of the coronavirus. Until the end of May, infection continued to spread mainly among adults and elderly people. This generation-specific interview survey of available nationwide data was conducted to ascertain infection sources and transmission in Japan, particularly to elucidate the actual circumstances of intergenerational infection and Japan’s strategic approach to respond to COVID-19.

## Methods

To conduct a retrospective observational case series study analyzing publicly available anonymous reports, we accessed information from local health authorities’ reports released for January 15, 2020 through July 31, 2020 in Japan. Cases were confirmed as SARS CoV-2 positive using RT-PCR testing by local health authorities, or using RT-PCR testing or other molecular testing with processing by commercial laboratories. Health center staff in each prefecture investigated the status of contact with each SARS CoV-2 infected person, identified the infection source, and reported it to their respective health authorities. The results are posted on the website of each prefecture and are accessible by anyone. To identify asymptomatic persons, it is necessary to examine all the members in clusters accurately. Invariably only a part of them can be acquired. Therefore, asymptomatic persons were excluded.

After excluding asymptomatic persons, we ascertained whether each symptomatic SARS-CoV-2-positive-confirmed person was a source of infection in a cluster at homes, schools, and daycare facilities during January 15, 2020 – July 31, 2020. Different distributions of sources of infection were analyzed by generation. Statistical analyses were applied using software (R ver.5). Using available information related to January 15, 2020 – April 24, 2020, we retrospectively identified cases in which a child transmitted the virus to other children in clusters at daycare facilities. Because this is a case series study and because information to use was available from the websites of all prefectures in Japan, no approval by an ethical committee was necessary.

For this study, *x*_*i,j*_ was defined as the number of patients in *i* age group who were infected from patients of the *j* age group. Particularly, *i*(*j*) *=* 1 represents an age class younger than 20 years old, 2 denotes the age group of patients who were 20 years old or elder than 20 years old and younger than 60 years old, and 3 stands for an age group of people older than 60 years old. Additionally, *i=*4 represents patients whose source of infection was unidentified.

The source of infection of most patients was unidentified. Therefore, we assumed that the age distribution of source of infection (*x*_*i,1*_,*x*_*i*,2_ and *x*_*i*,3_) was the same among patients whose source of infection was not identified (*x*_*i*,4_). Then, we arranged the Table as *y*_*i,j=*_ *x*_*i,j*_ *Σ* _*k=1*_^4^ *x* _*i,k*_ */Σ* _*k=1*_^3^ *x* _*i,k*_ for *i*(*j*)*=*1,2, and 3.

For this study, *R*_*i,j*_ which represents the effective reproduction number of the *j* age group infection to the *i* age group was defined as *y*_*i,j*_*/Σ*_*k=1*_^3^ *y*_*j,k*._ As one might expect, it was correct when *i* and *j* were not the same. However, if *i* and *j* were the same, patients were also probably the source of infection to patients of the same age group. Therefore, double counting occurred in the infected patients and source of infection. *R*_*i,i*_ cannot be defined correctly using this procedure.

The confidence interval (CI) of *R*_*i,j*_ was estimated using the following method. Because we do not know the probability that a patient infected one patient, the probability that a patient infected two patients, or more patients, we presumed that these were *p, p*^*2*^, *p*^*3*^, and so on. Then *R*_*i,j=*_ *p+*2*p*^*2*^*+*3*p*^*3*^*+*…*=Σ*_*k=*1_ *k p* ^*k*^*=p/*(1-*p*)^2^. Therefore, we can convert *R*_*i,j*_ to *p* uniquely. We denoted *p*_*i,j*_ as converted from *R*_*i,j*_. We estimated its CI through bootstrapping. We first draw *M=Σ* _*k=1*_^*3*^ *y* _*j,k*_ random numbers of the uniform distribution on (0,1) from the generator. We designate it as z_*m*_ (*m=*1,2,…*Σ*_*k=*1_^3^ *y*_*j,k*_). If z_*m*_ <*p*_*i,j*_, then the *m*-th patients infected more than one patient. Similarly, if z_*m*_ <*p* _*i,j*_^2^, then the *m*-th patient infected more than two patients, and so on. Therefore, *Σ*_*k=1*_*k* 1[*z*_*m*_ *< p*_*i,j*_*k*] was the estimated number of people infected by the *m*-th patient, where the function of 1[] was one if [] was true and zero otherwise. Similarly, *Σ*_*m=1*_^*M*^*Σ*_*k=1*_*k* 1[*z*_*m*_ *< p*_*i,j*_*k*] was one of the bootstrapped *R*_*i,j*_. We repeated this procedure one million times. Then we obtained the one million estimator of *R*_*i,j*_. We sorted these variables. The duration from the first 25 thousandth *R*_*i,j*_ to the last 25 thousandth *R*_*i*,_ should be CI of *R*_*i,j*_.

## Results

The status of SARS-CoV-2 outbreaks up to November 17, 2020 is presented in Figure 1. The period of school closures from March 2 to the end of May 2020 and that of the state of emergency when the Japanese government was asking people in Japan to stay home as much as possible to prevent the spread of the coronavirus is shown in Figure 2. Symptomatic and asymptomatic SARS-CoV-2-positive-confirmed persons are also shown in Figure 2, which shows a small number of people with symptoms and most people as asymptomatic and under investigation. The SARS-CoV-2 confirmed cases comprised those of three patient age groups: underage people under 20 years old; adults 20–59 years old; and elderly people 60 years old and older. Of the 26,986 cases of SARS-CoV-2 confirmed from January 14 through July 31, 2020, 3,240 were ascertained as sources of infection. Of them, 125 (3.9%) were of underage people, 2,350 (72.5%) were of adults, and 765 (23.6%) were of elderly people (Table 1). It was impossible to ascertain if the remaining 23,746 of the 26,986 cases were sources of infection or not. The overwhelming majority of SARS-CoV-2 infections therefore occurred among adults and elderly people.

**Table 1.**
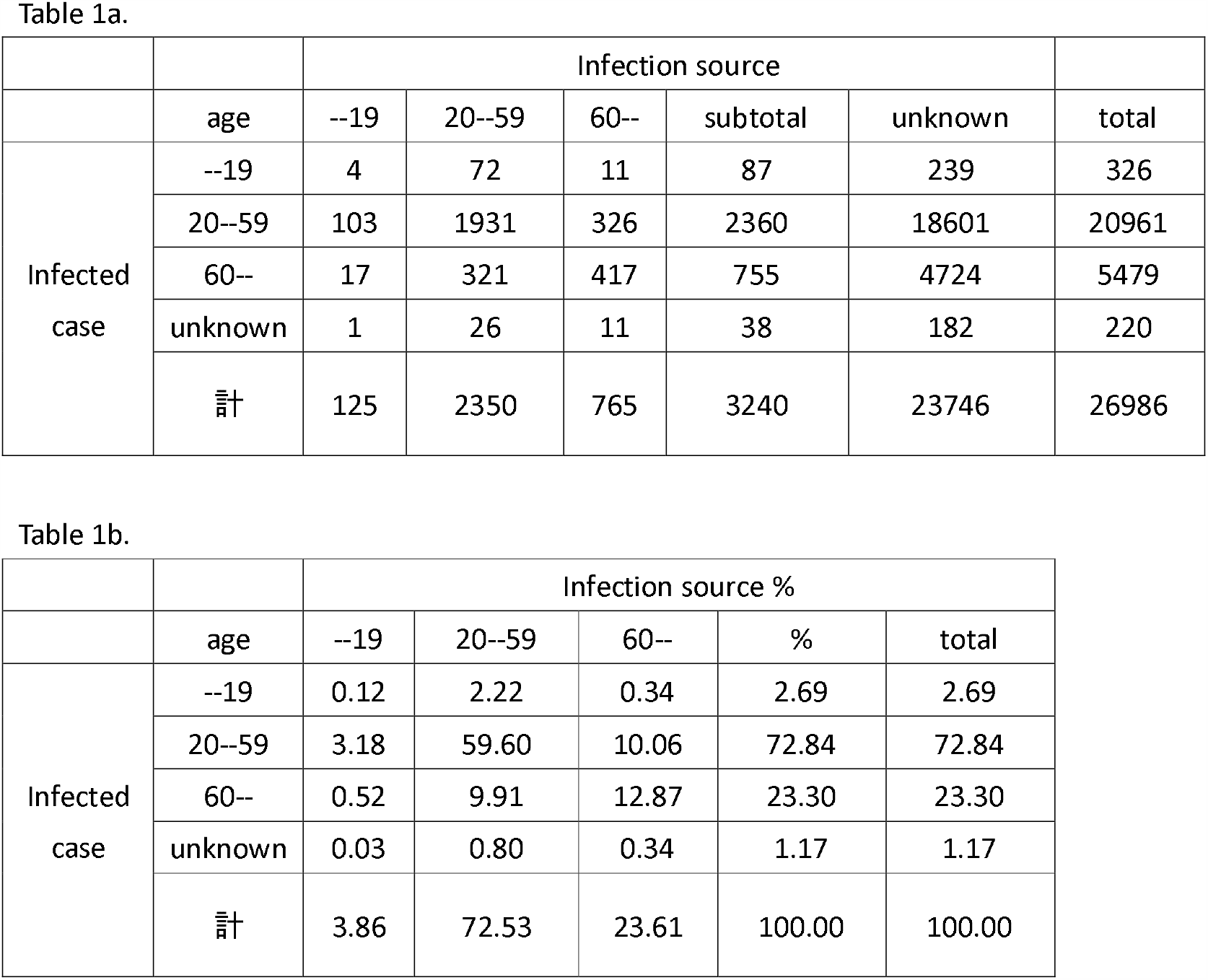
Age-dependent difference in distribution of SARS-CoV-2 infection sources among age groups

**Figure 1.**
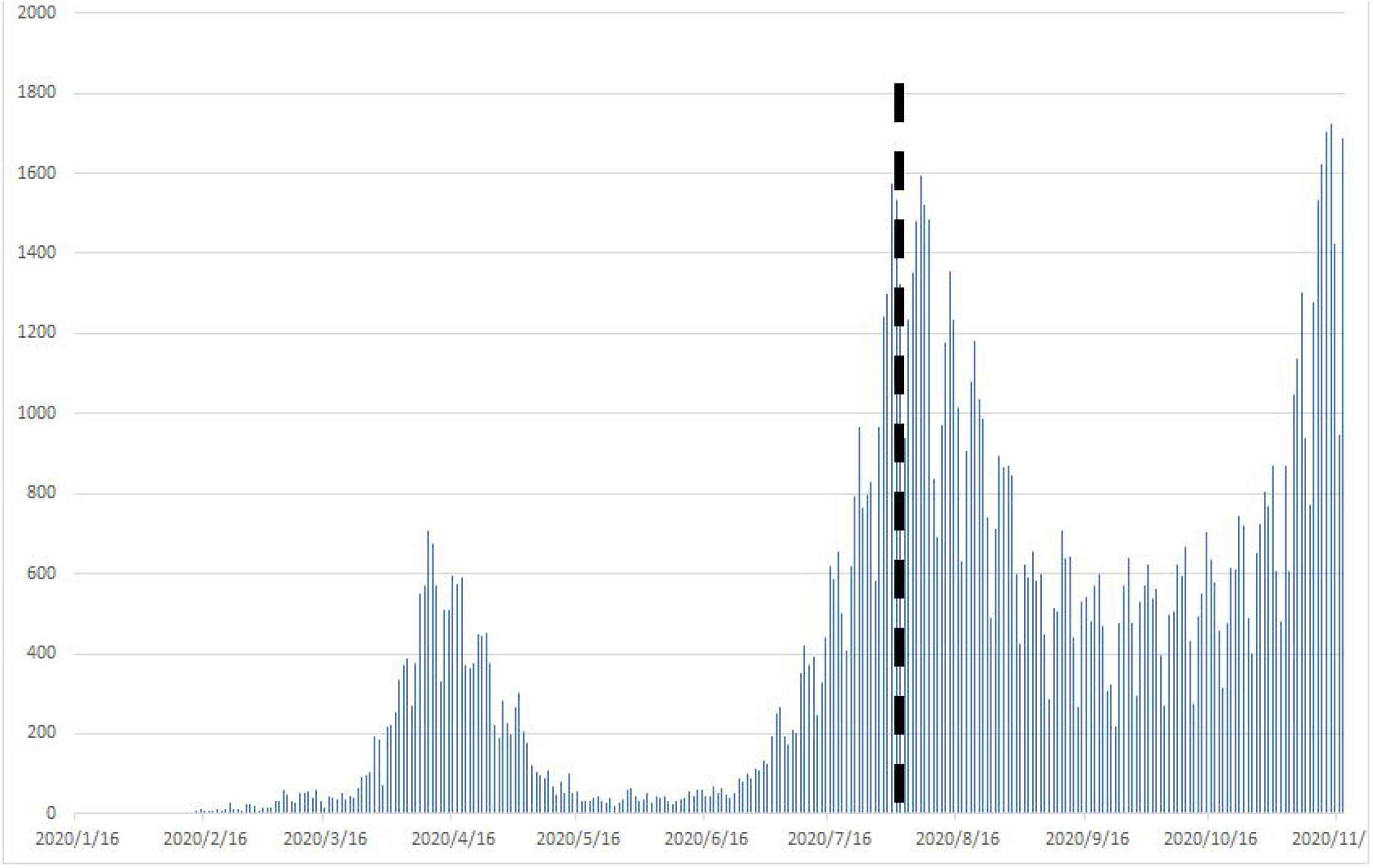
SARS-COV-2 outbreak during Jan. 14 – Nov. 17, 2020 in Japan.

**Figure 2.**
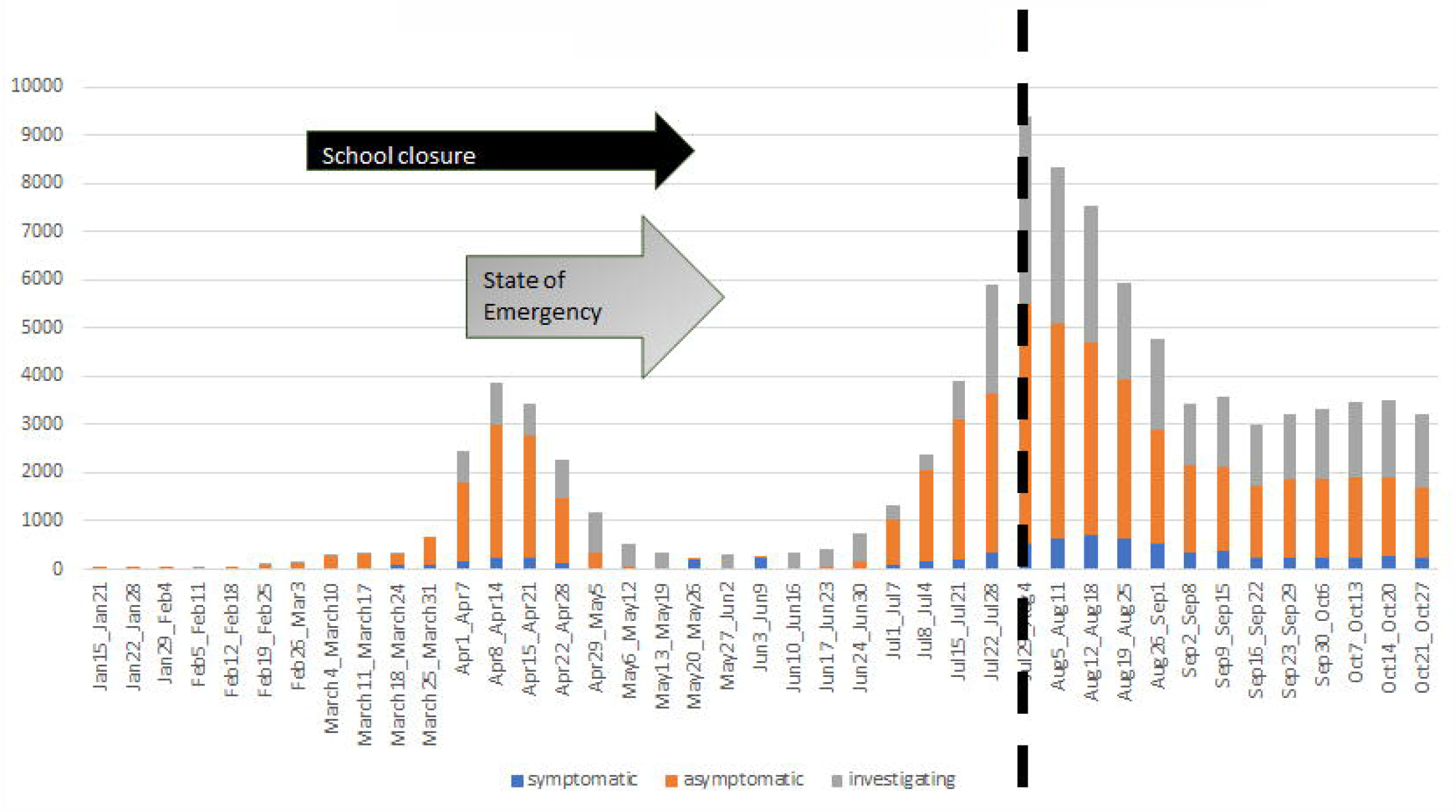
Dynamics of SARS-COV-2 infected people including various administrative measures from Jan. 14 – Oct. 27, 2020 in Japan.

Reproductive numbers (R) of sources of SARS-Co-V2 infection were estimated for three age groups (Table 2). The respective R numbers of underage sources of infection showing the direction of transmission to underage people, adult people, and elderly people, estimated as 0.0415 (95% CI, 0.0138–0.0691), 1.11 (95% CI, 0.9171–1.3226), and 0.2811 (95% CI, 0.2074–0.3687). The respective R numbers of adult source of infection showing the direction of transmission to underage people, adult people, and elderly people were estimated as 0.0140 (95% CI, 0.0120–0.0162), 0.5392 (95% CI, 0.5236–0.5550), and 0.1135 (95% CI, 0.1074–0.1197). The respective R numbers of elderly sources of infection showing the direction of transmission to underage people, adult people, and elderly people were estimated as 0.065 (95% CI, 0.0039–0.0091), 0.3264 (95% CI, 0.3059–0.3474), and 0.3991 (95% CI, 0.3757–0.4229). The respective numbers (percentages) of presumed areas of transmission were 1,109 (4.0) of families, 778 (2.8) of hospitals and clinics, 337 (1.2) of nursing homes, 24 (0.1) of schools excluding vocational schools and universities including kindergartens, 19 (0.1) of daycare facilities, 45 (0.2) of vocational schools and universities, 151 (0.5) of restaurants, 47 (0.2) of night entertainment, 37 (0.1) of karaoke, 68 (0.2) live theaters, 12 (0.04) of sports facilities, 538 (1.93) of companies and offices, 5,411 (19.41) of others, and 19,303 (69.24) of unknown places. Excluding the unclassified areas and unknown areas, family, hospitals and clinics, nursing homes, schools excluding vocational schools and universities including kindergartens, daycare facilities, vocational schools and universities, restaurants, night entertainment, karaoke, live theater, sports facilities, company/office were accounted 35, 25, 11, 1, 1, 1, 5, 1, 1, 17, 0, and 2%, respectively (Fig. 3b). Actually, 69.2% of all infected people were unable to identify the location of the infection. Of 11.35% of the identified sources of infection, homes, hospitals and nursing facilities, other activity places including schools, and workplaces accounted for 1/3 each of those identified locations (Fig. 3).

**Table 2.** Reproductive numbers of source of SARS-Co-V2 infection in 3 different Age groups

| age group of infected cases | R | Confidence interval |
| --- | --- | --- |
| --19 | 0.0415 | [0.0138, 0.0691] |
| 20--59 | 1.1106 | [0.9171, 1.3226] |
| 60-- | 0.2811 | [0.2074, 0.3687] |

2b. Infection source: 20-59 years old
| age group of infected cases | R | Confidence interval |
| --- | --- | --- |
| --19 | 0.0140 | [0.0120, 0.0162] |
| 20--59 | 0.5392 | [0.5236, 0.5550] |
| 60-- | 0.1135 | [0.1074, 0.1197] |

2c. Infection source: >59 years old
| age group of infected cases | R | Confidence interval |
| --- | --- | --- |
| --19 | 0.0065 | [0.0039, 0.0091] |
| 20--59 | 0.3264 | [0.3059, 0.3474] |
| 60-- | 0.3991 | [0.3757, 0.4229] |

**Figure 3a.**
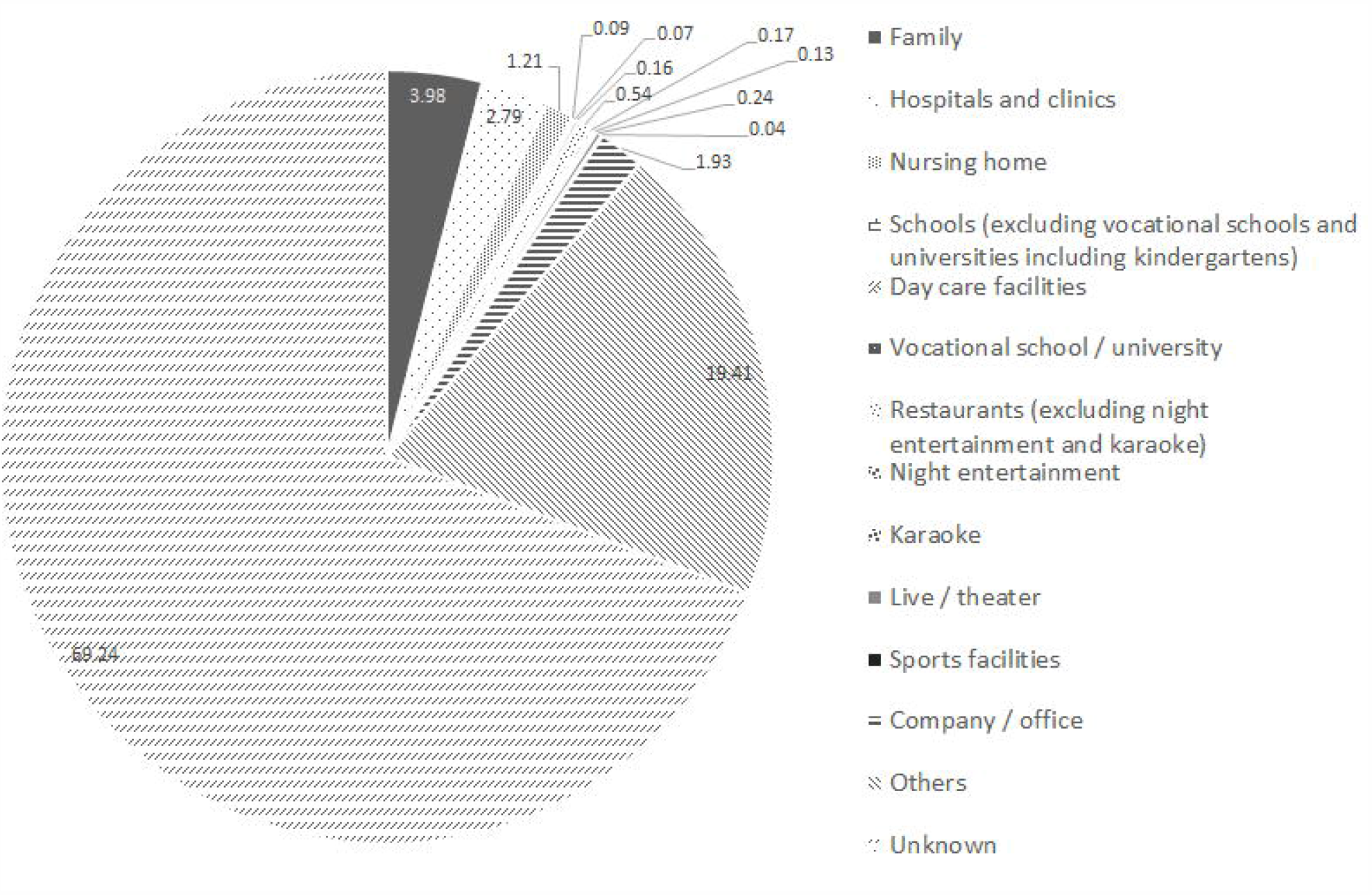
Estimated infection locations of SARS-COV-2 infected persons who became the infection source.

**Figure 3b.**
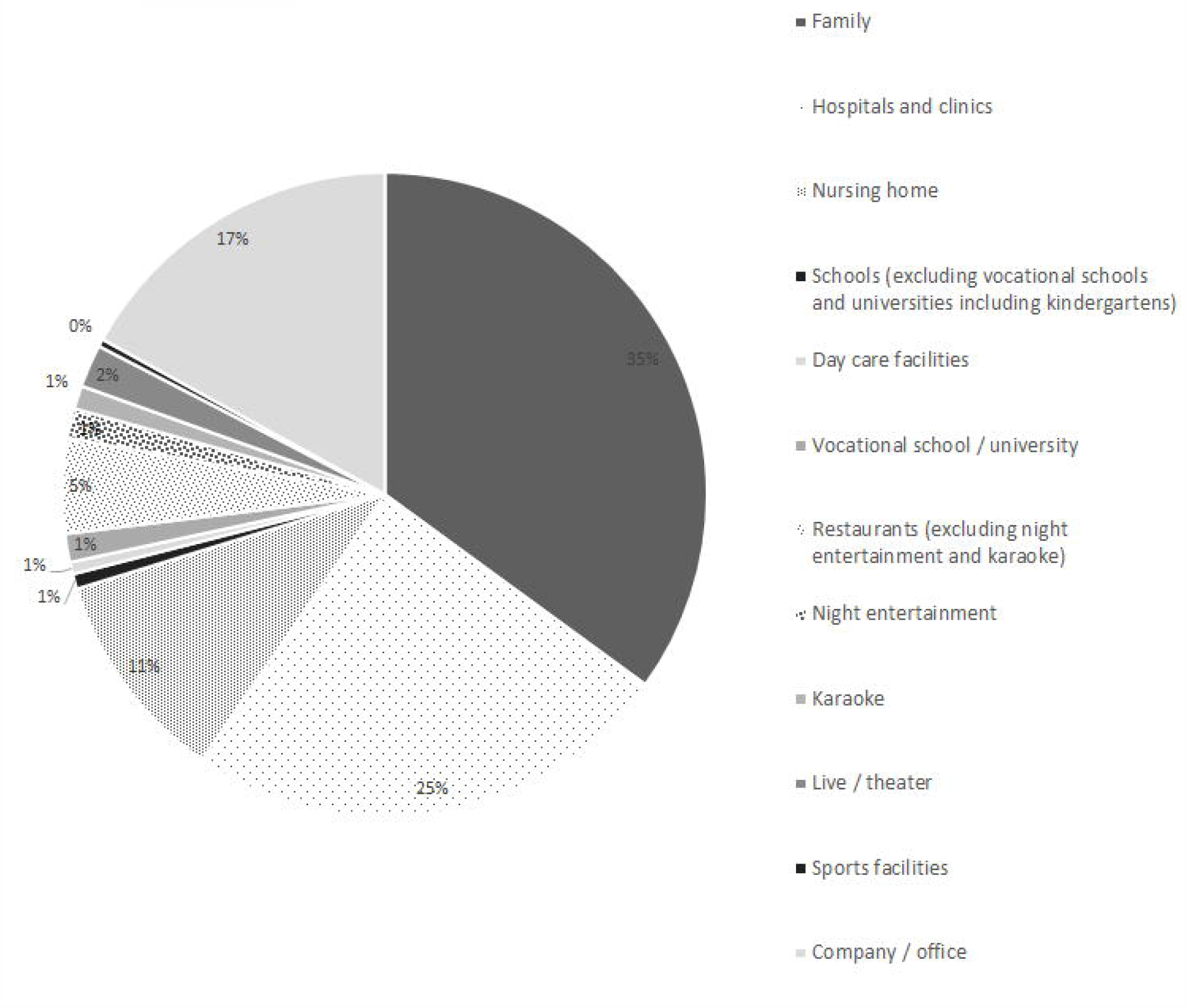
Rate of estimated infection locations as specified by SARS-COV-2 infected persons who became the infection source.

Table 2

Age-dependent difference in R number among three age groups

## Discussion

We investigated details of SARS-CoV-2 transmission modes in Japan. Data in this paper constitute a compilation of information of symptomatic SARS CoV-2 infected persons published by prefectures. No other summary regarding infection sources has been reported in Japan.

This report is the first of a study elucidating age-dependent distribution differences of infection sources for SARS-CoV-2 transmission in Japan. We assessed detailed descriptions in reports of symptomatic SARS-CoV-2 confirmed cases in schools, kindergartens and daycare facilities, companies and offices, etc. through July 31, 2020. The respective Rs of transmission from adults and elderly people were less than 1. The R from the underage group for adult patients confirmed to be SARS-CoV-2 positive was 1.11. Only this combination was found to be greater than 1. The R numbers in the transmission clusters for which we were able to identify the infection source were almost all less than 1.

The basic reproduction number R_0_ of SARS-CoV-2 was first estimated as around 2.2 (90% high density interval: 1.4–3.8) in China [4]. The R_0_ was estimated as 2.56 in Japan, with 95% CI of [2.51, 2.96] before school closure [5]. In the case of outbreak of COVID-19 in the Princess Cruise ship, an estimate of the mean reproduction number in the confined setting reached values as high as approximately 11 [6]. The highest reproductive number was reported for the Diamond Princess Cruise Ship in Japan (14.8); the estimated summary reproductive number was 2.87 (95% CI, 2.39–3.44 [7].

It is noteworthy that the R numbers of underage people as infection sources from the COVID-19 patients in all age groups were lower than those of adults and elderly people. Rarity of underage people of SARS-CoV-2 transmission was found. When we identified patients as sources of SARS-CoV-2 infection, hospitals, clinics and day care facilities, family members, and companies each accounted for approximately one-third of the infected area. Results show that the age distribution of SARS-CoV-2 infection sources reflects different social age-related activities. It became very difficult to ascertain the infection source and the age group of infected persons because of the dramatic increase in infection in adults. It remained in the very early stages of the pandemic. Unfortunately, it was not possible to ascertain the infection source and the age group of the infected person by type of labor during this pandemic. It might be possible to do it after the end of the pandemic in the future, but it can not be done in time as a countermeasure for the ongoing epidemic. Therefore, the purpose for this study was to reach a certain conclusion for an analyzable research period.

The Japan Pediatric Society reported the Current Status of Medical Findings on Novel Coronavirus Infections in Children on May 7, 2020. Findings up to the present day for COVID-19 indicate that it is unlikely to spread in clusters at schools and at group care facilities, as it does for influenza which is a severe disease that mainly affects children [8]. In our study, only four probands under the age of 20 were the source of infection for persons under the age of 20 with confirmed SARS-CoV-2 positivity (Table 1a). Although data for positive cases in schools are limited, they have not demonstrated a remarkable degree of transmission. In one pediatric case, a child visited three schools while symptomatic, but did not transmit the virus to any of 112 people who had close contact with him, suggesting that children might not be an important source of virus transmission [9]. Gradually, it has been clarified that children can escape SARS COV-2 infection well [10, 11].

Children mainly transmitted SARS-CoV-2 to family members. In fact, from a study using epidemiologic data, 95.59% (65 of 68) of infected children were found to have been infected through household contact with adults [12]. A study of the dynamics of SARS-CoV-2 transmission within families has indicated that children are less likely to show SARS-CoV-2 positive results than coresident adults are [13]. In addition to this evidence, our study of cases and their infected and uninfected close contacts provide key insights supporting epidemiologic studies of SARS-CoV-2. Household contacts were the main transmission mode including children, but children rarely spread the virus to other children and teachers at schools, kindergartens, and nursery schools. The main source of infection in society is not children, but adults or senior patients with SARS-CoV-2 infection. Illness and mortality rate data vary from country to country. Therefore, policies can also be expected to vary. However, because this infection is mainly an adult disease, infection control of this virus should be conducted specifically by generation. SARS-CoV-2 infection among children is apparently controlled sufficiently for school settings. Additional studies should be conducted to ascertain if school life can be undertaken safely as schools open gradually. Although schools in Japan have opened in earnest, household transmission was identified as a dominant route of students [14]. We hope that policies will be chosen based on carefully analyzed data from epidemiological surveys.

This study presents several important limitations. The most important limitation of this paper is that it analyzes only symptomatic individuals. This is true because, to study asymptomatic persons around the infected person, it is necessary to examine all the contact persons of the infected person, so that only a part of the relation is invariably acquired. As a result, the proportion of infected individuals is significantly lower than that of symptomatic individuals. A simple sum of the two does not make sense. However, it has been suggested that SARS Co-V2 is highly infectious even among asymptomatic individuals. It cannot be ruled out that the asymptomatic epidemic might spread in schools or nurseries if the proportion of asymptomatic children is high and their infectivity is high. To confirm this point, continuous inspection of all children and children at school or nursery school is necessary. No such design findings in the future have been published at this time. Apparently, future consideration is necessary. Second, the data we used were based solely on information obtained through interviews by health center staff. Necessarily, not all information was disclosed. Analyses were conducted as well as possible given the limited data which were obtained. However, there were 506 unknown cases other than the 341 determined cases as sources of infection. Moreover, we did not measure viral loads in COVID-19 patients in these transmission clusters. Further studies should be conducted based on data accumulated during a longer period until the end of the second or third wave of imported COVID-19 cases in Japan. Third, because of widespread school closures during this study, most opportunities for infection among children were avoided in the first place, which might have contributed to the results. Of course, it cannot be denied that the chances of infection among children might have decreased because of school closures and closures of kindergartens. However, contact opportunities at home do not change because of these closures, or by refraining from going out. Rather, the frequency of contact over the ages would probably increase. In addition, contact opportunities outside of schools, kindergartens, and nursery schools were not eliminated completely. It is noteworthy that the examination in this paper under such circumstances is an interpretation. Therefore, the conclusions in this paper will not actually hold true when schools or nurseries are reopened. However, the virus is expected to be spread mainly among adults.

Although the only R higher than 1 was found to be that of underage people showing the direction of transmission to adults, underage people constituted only a minority of infected cases. Adults and elderly people were widely infected with SARS-CoV-2. The main source of SARS-CoV-2 infection is not children, but rather adults and elderly people infected with SARS-CoV-2. Establishment of new age-dependent isolation strategies to avoid spreading the viral infection are highly anticipated. Because adults transmit the disease easily to humans, if an adult infected person is found, an adult close contact who is likely to become severely infected should be examined. If infected, an isolation policy is taken. However, children are less likely to become severely ill and are less likely to transmit the virus to similarly underaged people and elderly people. For that reason, their isolation policy warrants some consideration. Only children with close contact to an infected underage person with SARS-CoV-2 should be isolated and inspected. Further studies are now being conducted based on data from COVID-19 cases accumulated over a longer period of time in Japan.

## Data Availability

Data used in the paper are publicly available.

https://www.mhlw.go.jp/stf/seisakunitsuite/bunya/0000164708_00001.html#kokunai2hassei.

## Acknowledgments

We are grateful to all patients who contributed to this study and to their families, as well as staff of public health centers nationwide, who helped collect patient information.

## Conflict of Interest Disclosure (includes financial disclosure)

No author has any conflict of interest to disclose.

## Funding/Support

This research benefited from no specific grant from any funding agency in the public, commercial, or not-for-profit sector.

## Notes

### Competing Interest Statement

The authors have declared no competing interest.

### Author Declarations

Ethical approval was not requred as this research is based on secondary analysis of publicly available data.

